# Evaluation of pre-Omicron population immunity for SARS-CoV-2 in the Household Influenza Vaccine Evaluation (HIVE) cohort

**DOI:** 10.64898/2026.03.10.26348071

**Authors:** Yangyupei Yang, Casey Juntila, Amy Callear, Matthew Smith, Reilly K. Atkinson, Yan Zhao, Ruhi Shah, Jefferson J. S. Santos, Scott E. Hensley, Arnold S. Monto, Emily T. Martin

**Affiliations:** Department of Epidemiology, University of Michigan School of Public Health, Ann Arbor, MI, USA; Department of Microbiology, Perelman School of Medicine, University of Pennsylvania, Philadelphia, PA, USA

**Keywords:** COVID-19, SARS-CoV-2, antibody, immunity, immunoassay

## Abstract

**Background:** Continued serosurvey for SARS-CoV-2 population immunity is critical for vaccine strain selection and identification of susceptible groups such as young children. We aim to evaluate the use of a high-throughput, low-volume assay to describe strain-specific IgG measurement in a longitudinally followed community-based cohort.

**Methods:** The longitudinal Household Influenza Vaccine Evaluation study has observed around 300 households annually for acute respiratory illness. SARS-CoV-2 variant-specific IgG (including original, delta, beta, and omicron variants) concentrations were measured using multiplex electrochemiluminescence (ECL), pseudoviral neutralization (PN), and ACE2 inhibition assays in participants who had serum samples drawn between July 1 and December 31, 2021. Spearman’s rank correlation coefficient assessed correlation within and between ECL, ACE2, and PN assays. Receiver operating characteristic (ROC) analysis evaluated the performance of ECL and ACE2 assays. Generalized Additive Mixed Models estimated population immunity by age against SARS-CoV-2 variants.

**Results:** A total of 156 serum samples from 117 participants were selected for this study. Most were from adults and a majority received vaccination during the analysis period. High correlations were observed within ECL (0.99-1.0), ACE2 (0.87-0.99), and between panels: ECL vs. PN (0.95-1.00), ACE2 vs. PN (0.85-0.96). ROC analysis showed medium to high areas under the curve, sensitivity, and specificity for ECL (0.85-0.87; 0.89-0.90; 0.87-0.88) and ACE2 (0.85-0.96; 0.94-1.0; 0.42-0.92). SARS-CoV-2 IgG concentrations are higher in older individuals, plateauing at age 20.

**Conclusion:** We found a high correlation between ECL, ACE2, and PN across variants. Age-specific IgG quantities reflected behavior and infection histories collected by active surveillance.

## Background

COVID-19, caused by the SARS-CoV-2 virus, has shown a significant health burden globally, characterized by its rapid evolution and severe disease outcomes^1^. The immunoglobulin G (IgG) generated either post-vaccination or through natural infection is pivotal in preventing severe outcomes and, to some extent, subsequent infections^2^. Cross-protection has been observed where IgG responses to one strain may cross-protect against other strains^3^. Serological assays, which quantify specific immunoglobulins such as IgG neutralizing or functional antibodies, are important for understanding population immunity. Pseudovirus neutralization (PN) assays, a functional test that requires blocking viral entry and replication, typically focus on a single protein target per test and are technically challenging^4^. The complexity of immune responses to fast-evolving SARS-CoV-2 strains needs a broader approach. Multiplex electrochemiluminescence (ECL) assays assess antibody binding and can simultaneously detect multiple antigens. However, concerns remain that binding assays may not adequately assess the capacity to block infection and could potentially lack specificity against different variants. Another competition multiplex assay evaluates antibodies’ capacity to inhibit the binding of the angiotensin-converting enzyme 2 (ACE2) to SARS-CoV-2 antigen, thus analyzing antibody activity through ACE2 binding inhibition. Multiplex assays provide a robust and scalable solution for comprehensive immune profiling that can be used for large population-based serosuveys^5,6^.

We conducted a multi-age community serosurvey of pre-Omicron antibody levels using a multiplex coronavirus immunoassay from Meso Scale Diagnostics (MSD, Rockville, USA) that utilizes electrochemiluminescence for the detection of antibodies against various protein targets, including the Spike protein, the Receptor Binding Domain (RBD) of the spike protein, and the nucleocapsid protein (N) of several SARS-CoV-2 strains. Additionally, our study used the ACE2 multiplex assay for antibodies that block the binding of ACE2 to Spike antigens from variants of SARS-CoV-2 to assess binding inhibition against various SARS-CoV-2 antigens. These assays were compared to PN tests, providing a comprehensive evaluation of the baseline antibody profile of participants in the longitudinal Household Influenza Vaccine Evaluation (HIVE) cohort conducted in Michigan before the spread of the Omicron variant. A major strength of the longitudinal HIVE cohort is the availability of repeated, high-quality immunologic measurements that capture longitudinal antibody immunity along with complete vaccination and infection history, making this a unique resource for nested serosurveys^7–12^.

The period before widespread Omicron transmission (July 1^st^ – December 31^st^ 2021) offers a unique opportunity to study SARS-CoV-2 infection- and vaccine-induced immunity, as infection prevalence was low compared to the Omicron period, and vaccines had not yet been introduced to children. This created a clear distinction between infection-induced and vaccine-induced immunity, with few cases of infection in vaccinated individuals (i.e. “hybrid immunity”).

## Method

### 2.1 Study population and serum samples

Recruitment and retention procedures of HIVE study have been previously described^13^. Briefly, eligible households must have ≥3 persons living at the same address with at least one child aged <10 years at the time of initial enrollment. In mid-2020, eligibility was expanded to those with at least one child aged <21□years. Participants contributed blood specimens at enrollment, twice annually, pre- and post-flu and COVID-19 vaccination. For this analysis, serological samples were selected among HIVE participants who provided blood specimens between July 1^st^ and December 31^st,^ 2021.

### 2.2 Demographic Information and Household Characteristics

Each participant, or their parent or legal guardian, completed an enrollment survey detailing demographics, household characteristics, chronic medical conditions, COVID-19 vaccination history, and prior SARS-CoV-2 infection. Participants were surveyed annually to update demographic and household information.

Weekly surveillance for COVID-19 symptoms was conducted. Households received weekly email reminders to report acute respiratory illnesses (ARIs) (see Supplementary Materials) at symptom onset (within 7 days post-onset) of qualifying ARIs by parents or through self-collection. All respiratory specimens were tested for SARS-CoV-2 using multiplex reverse transcriptase-polymerase chain reaction (RT-PCR).

Pre-Omicron Individual immunity profiles were classified into four categories: 1) Vaccination: participants who received at least the primary doses of COVID-19 vaccination as defined by the CDC; 2) Infection: participants with RT-PCR confirmed SARS-CoV-2 symptomatic infection before the emergence of Omicron; 3) Hybrid Immunity: participants who received at least the primary doses of COVID-19 vaccination and had RT-PCR confirmed SARS-CoV-2 infection before Omicron; 4) No vaccination, no infection.

### 2.3 Serological assays

Serum specimens were screened for IgG antibodies to the SARS-CoV-2 spike, RBD, and N protein, using a commercially available electrochemiluminescence (ECL) immunoassay kit Panel 29 (Spike antigens: original, B.1.351, B.1.617.2, BA.2, BA.2.12.1, BA.2.75, BA.4, BA.5)) and 30 (RBD antigens: original, B.1.1.7 B.1.351, BA.2, BA.2.12.1, BA.2.75, BA.4/5, delta. A result of 5000AU/mL N protein was considered seropositive^14^. Serum samples were diluted 1:5000 and 1:50,000 and run in duplicate on each plate and processed following the manufacturer’s protocol^15–17^. Two serology controls (SC 1.2 and SC 1.3) provided by the manufacturer, containing known concentrations of IgG antibodies against the targets in the panel, were run in duplicate on each plate. Four independent runs for each sample were used to calculate coefficient of variation (CV) for quality control. Assay data were processed using standardized R code to resolve duplicates and reflex testing to higher dilutions (supplementary material).

Competition ACE2 assays were used to evaluate the antibody inhibition capacity against SARS-CoV-2 antigens, specifically targeting spike and RBD proteins^18,19^. The percentage inhibition was calculated relative to the assay calibrator (maximum 100% inhibition) using the equation:

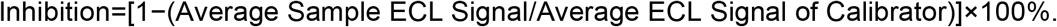

PN assays were conducted to measure serum titers of neutralizing antibodies against Wild Type (WT) and BA.1 Spike strains, as previously described^20,21^. Briefly, VeroE6/TMPRSS2 cells were seeded at 2.5×10^4^ cells/well in collagen-coated 96-well plates overnight. The following day, heat-inactivated sera were serially diluted 2-fold and mixed with ∼300 focus-forming units per well of each VSVΔG-RFP SARS-CoV-2 pseudotype virus. A mouse anti-VSV G monoclonal antibody, 1E9F9, was spiked into virus-antibody mixtures at 600 ng/mL to ensure neutralization of any VSV-G carryover virus from pseudotype virus production. Virus-antibody mixtures were incubated for 1 h at 37 °C and then transferred to VeroE6/TMPRSS2 cells. After 22 hours postinfection at 37°C, cells were washed with PBS, fixed with 4% PFA, and then blotted dry. Fluorescent foci were visualized and counted on an ImmunoSpot S6 analyzer. The focus reduction neutralization titer 50% (FRNT50) was measured as the greatest serum dilution at which the foci count was reduced by at least 50% relative to control cells that were infected with pseudotype virus in the absence of participant serum. FRNT50 titers for each sample were measured in two replicates.

### 2.4 Statistical analysis

Antibody concentrations were analyzed for individual spike, RBD, and nucleocapsid (N) antigens measured on the ECL panels, as well as PN titers against wild-type and BA.1 spike. Seropositivity for N was defined using the manufacturer-provided cutoff 5000AU/mL by ECL assay. Spearman correlation was used to compare antibody levels within and between ECL panel variants, and to compare between PN and ECL assays. Geometric mean concentration (GMC) and 95% confidence interval (95%CI) were used to assess the GMCs of antibody concentrations for each strain among age categories. P values <0.05 were considered significant. A receiver operating characteristic (ROC) curve analysis was performed to evaluate the ECL and ACE2 assay performance by antigen and protein targets. ROC curves were generated, and the area under the curve (AUC) was calculated to assess discrimination. The optimal classification threshold was determined using Youden’s Index, which is the point on the receiver operating curve where sensitivity plus specificity is maximal, and aims to select an optimal threshold value for a diagnostic test ^22^. At this threshold, sensitivity, specificity, and their 95% confidence intervals (CIs) were calculated using the Wilson score method. A generalized additive mixed model was used to fit antibody trends by age and SARS-CoV-2 strains pre-Omicron. All analyses were conducted using R version 4.0.38^23^, and plots were generated using the ggplot2 package.

## Results

### 3.1 Participants and samples

Among the participants enrolled in the HIVE cohort, 156 serum samples from 117 participants were selected for this study (Table 1). The age group contributing the most serum samples (n=77, 66%) was over 18 years old. More eligible samples were tested from female participants (n=69, 59%). Among those 18 years and older, the majority (n=75, 97%) received vaccination (61 received vaccination only, 14 had hybrid immunity); 2.6% (n=1) were unvaccinated and uninfected during the analysis period. Whereas, among those younger than 18 years, 54% (n=21) were unvaccinated and uninfected, 7.7% (n=3) were infected only. The majority of participants were white (n=87, 75%), non-Hispanic (n=112, 97%), and 35% having high-risk conditions.

**Table 1:**
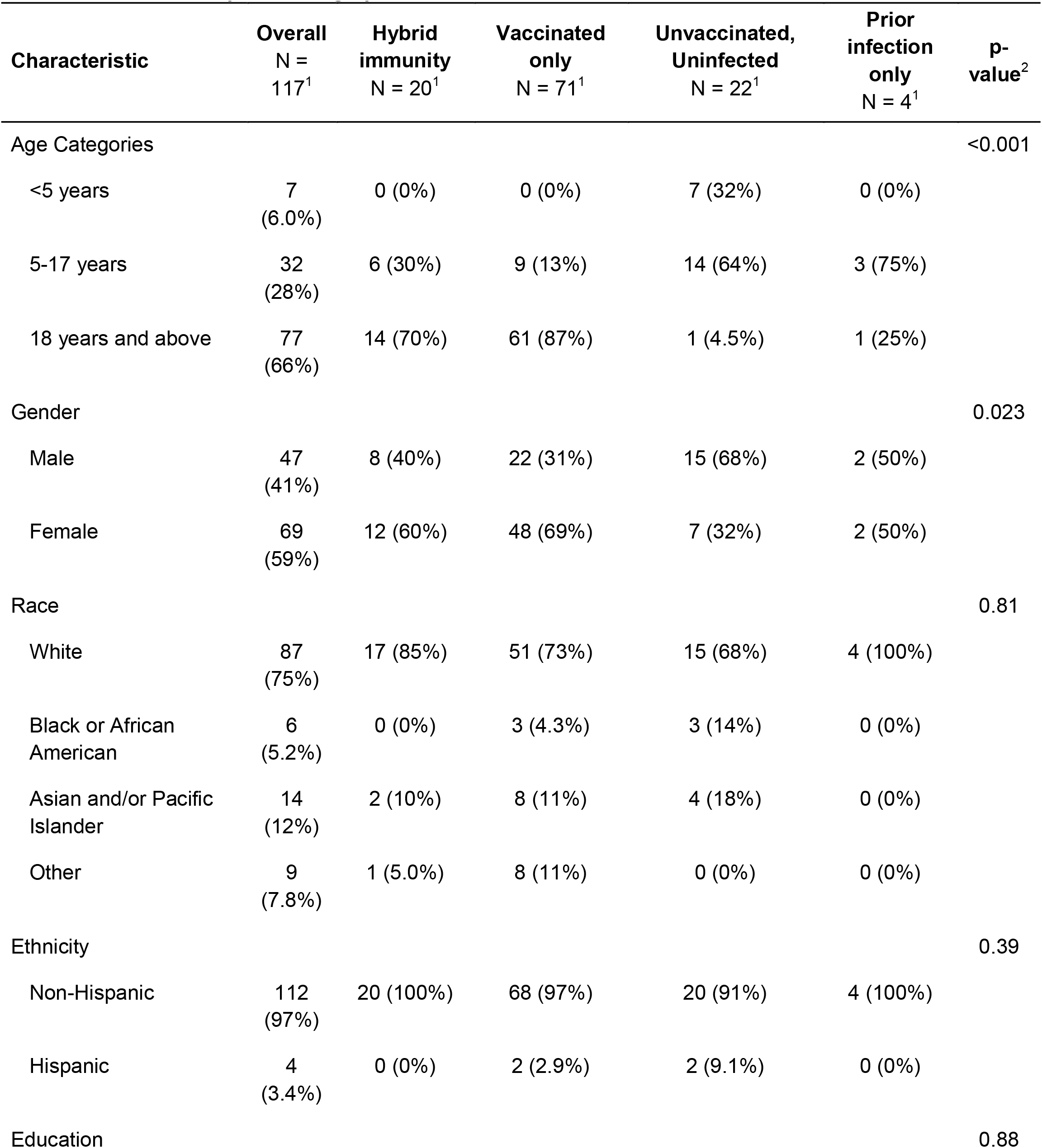
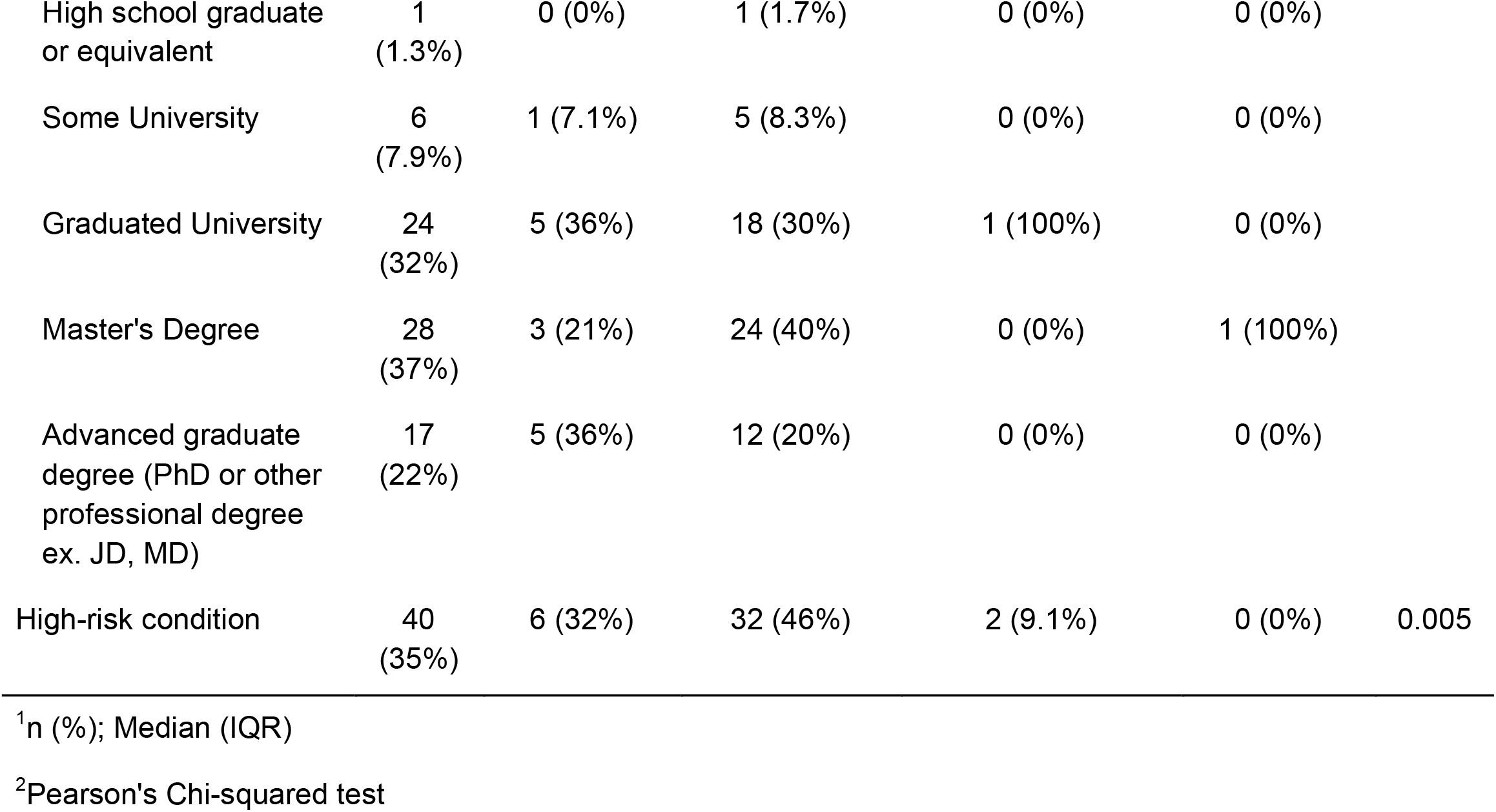
Participant demographic characteristics.

### 3.2 Comparison of multiplex electrochemiluminescence and pseudo-neutralization

Correlations between ECL measured IgG concentrations against Spike and RBD antigens were high across all strains, ranging from 0.954 to 0.996 (p<0.05) (Figure S1). Cross-assay correlations were evaluated for matched variants only. Specifically, ECL spike and RBD IgG concentrations were compared with PN titers for the same variant, and correlations were moderately high but lower than within-panel ECL correlations for both Spike and RBD panels, ranging from 0.811 to 0.827. Spike and RBD ACE2 inhibition showed high correlation with PN titers for matched variants, ranging from 0.877 to 0.953. At comparable PN titer levels, ECL IgG concentrations were higher for the B.1.351 (Beta) and B.1.617.2 (Delta) variants compared to other variants.

### 3.3 Antibody concentration relationship between antigens

Among 156 serum samples, 17 (10.9%) were N-positive. Based on historical surveillance swab results in the cohort, 8 individuals (5.1%) had previously tested PCR-positive for SARS-CoV-2 prior to the time of serum collection, while 148 (94.9%) had no documented prior PCR-positive result. Among individuals with a prior PCR-confirmed infection, 6 (75%) were also N-positive. An additional 9 samples were N-positive without a recorded prior PCR-positive result, indicating that serological testing may capture prior infections that were not detected by PCR surveillance, including asymptomatic infections, as well as potential pre-2020 N protein cross-reactivity from other seasonal coronaviruses.

Pearson correlation coefficients were high across all antigen pairs: 0.991–0.999 (p<0.05) for Spike IgG, 0.974–0.999 (p<0.05) for RBD IgG, 0.866–0.992 (p<0.05) for Spike ACE2, and 0.701–0.990 (p<0.05) for RBD ACE2. Correlations between BA.2 and other variants were lower in ACE2 inhibition assays: 0.436–0.714 for Spike ACE2 and 0.329–0.435 for RBD ACE2 (Figure S1). These results indicate small heterogeneity in antibody cross-reactivity across variants and assay types.

### 3.4 Assay sensitivity and specificity

ROC curve analysis was conducted to evaluate the trade-off between sensitivity and specificity for each antigen by vaccination status. Among vaccinated participants, IgG assays showed high area under the curve (AUC) values for spike (0.85–0.87) and RBD (0.85–0.87), while ACE-2 assays showed moderate to high AUCs for spike (0.85–0.96) and RBD (0.53–0.96), except for BA.2 sublineages (Table S1). Thresholds for sensitivity and specificity were determined using Youden’s Index and are summarized in Table S1. Among vaccinated individuals, IgG assays demonstrated high sensitivity and specificity, indicating good ability to correctly identify those with and without prior vaccination (sensitivity: 0.89–0.91; specificity: 0.87–0.88; Table S1). ACE2 assays showed consistently high specificity but variable sensitivity, particularly with lower sensitivity for BA.2 sublineages (sensitivity: 0.43–0.92; specificity: 0.91–1.00; Table S1).

### 3.4 Pre-Omicron population antibody characteristics against SARS-CoV-2

SARS-CoV-2 ECL IgG concentrations were higher with increasing years of age and plateaued in individuals around age 20 across different antigens (Figure 1A). This pattern was consistent regardless of the specific antigen, indicating high cross-reactivity within the panel. Antibody concentrations in unvaccinated and uninfected participants remained consistently low compared to the other three categories across all variants and panels. Within each age group (<5 years, 5-17 years, 18 years and above), the GMC of IgG antibodies against SARS-CoV-2 Spike proteins showed substantial differences. For children under 5 years, GMCs ranged from 43 AU (95% CI: 5.4-333) [BA.2.75] to 78 AU (95% CI: 7.1-852) [Original strain]. For children aged 5–17 years, GMCs ranged from 626 AU (95% CI: 175-2,233) [BA.2.75] to 2,743 AU (95% CI: 629-11,958) [B.1.617.2]. And in adults older than 18 years, GMCs ranged from 115,117 AU (95% CI: 69,347-191,098) [B.1.351] to 192086 AU (95% CI: 126,210-292,348) [Original strain]. PN assay results showed patterns similar to the ECL IgG results for the original SARS-CoV-2 strain and the BA.1 variant, with antibody responses increasing with age and reaching a plateau in early adulthood (Figure1B).

A similar trend was observed for ACE2 assay results against the original SARS-CoV-2 strain, where percent inhibition increased with age and plateaued around age 20 (Figure 1C). However, patterns varied among different antigens in adults. Percent inhibition was highest for the original strain (81.7%, 95% CI: 79.2%–84.3%), followed by higher values for the B.1.351 variant (58.5%, 95% CI: 55.2%–61.7%) and the Delta (B.1.617.2) variant (73.2%, 95% CI: 70.3%–76.0%). In contrast, percent inhibition was lower for other BA variants, with mean inhibition ranging from 22.7% for BA.2 to 41.5% for BA.4. Results in the prior infection alone group showed a lower percent inhibition, more similar to unvaccinated, uninfected participants across antigens, which differs from the patterns observed in ECL results. When comparing the original strain to other antigens, percent inhibition exhibited a wide range (0%–100%) in adults for non-original antigens. This finding suggests stronger cross-reactivity between the vaccine strain (original), B.1.351, and Delta variants, but less pronounced cross-reactivity between the original strain and later-emerging BA variants.

## Discussion

This study offers a comprehensive characterization of pre-Omicron antibody levels using multiple serological assessment tools in a longitudinal household cohort population with known COVID-19 vaccination and infection history. Across the cohort, we observed high antibody concentrations in vaccinated individuals and cross-reactivity across SARS-CoV-2 variants. Correlations between assays varied by exposure history and assay type, with electrochemiluminescence (ECL) binding assays showing strong internal consistency and pseudovirus neutralization (PN) assays providing more accurate functional results. We found that pre-Omicron antibodies were increasing with age until about 20 years, regardless of target antigen and testing method. Beyond early adulthood, responses plateaued with an apparent dip around age 40. This dip most likely reflects timing rather than biology: adolescents and young adults often received vaccination closer to the serology draw, while many in their forties had a longer interval since the last vaccine or infection, resulting in lower recent antibody concentration. Second, the majority of our cohort participants are middle-aged with more heterogeneous exposure histories and product mixes, which can widen variance and pull smoothed estimates downward. Finally, the flexible spline in the model may reflect local fluctuations when sample sizes are small.

When stratifying by vaccination and infection history, correlations between antibody measurements were generally higher among vaccinated individuals. In contrast, correlation coefficients among previously infected individuals were more variable, particularly for ACE2 inhibition assays. Some BA.2 comparisons showed near-zero or slightly negative correlations when compared to original, delta, beta, and other omicron variants, which could result from either unique antigenic differences for BA.2, or greater uncertainty in assessing an antigenically drifted strain. Within each panel, antibody concentrations across different SARS-CoV-2 strains were highly correlated, reflecting high cross-reactivity. This pattern likely reflects recognition of conserved epitopes across variants or the presence of broadly binding but non-neutralizing antibodies^3^.

We also assessed the nucleocapsid (N) antibody results to identify potential asymptomatic infections not captured through PCR testing. Although the cohort conducted active surveillance for respiratory illness with systematic PCR testing of symptomatic illness, several participants who tested negative by PCR were seropositive for N, suggesting undetected prior infection. This finding indicates that even within a well-characterized cohort with active surveillance, some infections may not be captured through PCR-based case ascertainment, likely due to asymptomatic infections, mild illness that did not prompt swab collection, or infections occurring outside the surveillance period. N-based serology therefore provides complementary information for identifying prior infection history in longitudinal cohort studies. While N antibodies may wane over time, their presence remains a useful marker for differentiating infection-induced responses from vaccine-induced ones, especially in populations with low testing coverage or limited symptom reporting.

The strengths of this study include the use of multiple validated assay types, stratification by exposure status, and detailed evaluation of cross-variant immune responses. Limitations include small sample sizes within a limited period (pre-Omicron) in certain subgroups, particularly those with hybrid immunity, and potential under-detection of asymptomatic infections. However, the low antibody levels observed in some participants support the accurate classification of immune status. Future improvements to multiplex platforms, including enhanced variant-specific detection and reduced cross-reactivity, will further expand their value.

In conclusion, our results from this serosurvey nested within a prospective longitudinal household cohort provide important context for interpreting future cross-sectional population serosurveys, which lack detailed information on prior infection and vaccination history.

## Data Availability

All data associated with the study will be available upon request. All original code is deposited at GitHub: https://github.com/Jadeyyyp/MSD-data-cleaning- and is publicly available.

## Author Contributions

Conceptualization: Y.Y., E.T.M.

Lab testing: C.J., R.A., Y.Z., R.S.

Project Administration: A.C.

Data Curation: A.C., M.S., Y.Y.

Analysis: Y.Y.

Visualization: Y.Y., A.C.

Original draft writing: Y.Y.

Review and editing: E.T.H., C.J., A.C., M.S., S.E.H, J.J.S.S., A.S.M.

Funding acquisition: S.E.H., A.S.M., E.T.M.

Supervision: S.E.H., E.T.M.

## Data Availability Statement

No material has been used from other sources. All data associated with the study will be available upon request.

## Ethics Approval Statement

The University of Michigan Institutional Review Board gave ethical approval for this work.

## Patient consent statement

All patients consented to participate in the study.

## Acknowledgements

We would like to thank all individuals for their participation in the HIVE study.

## Data and code availability

**Figure 2:**
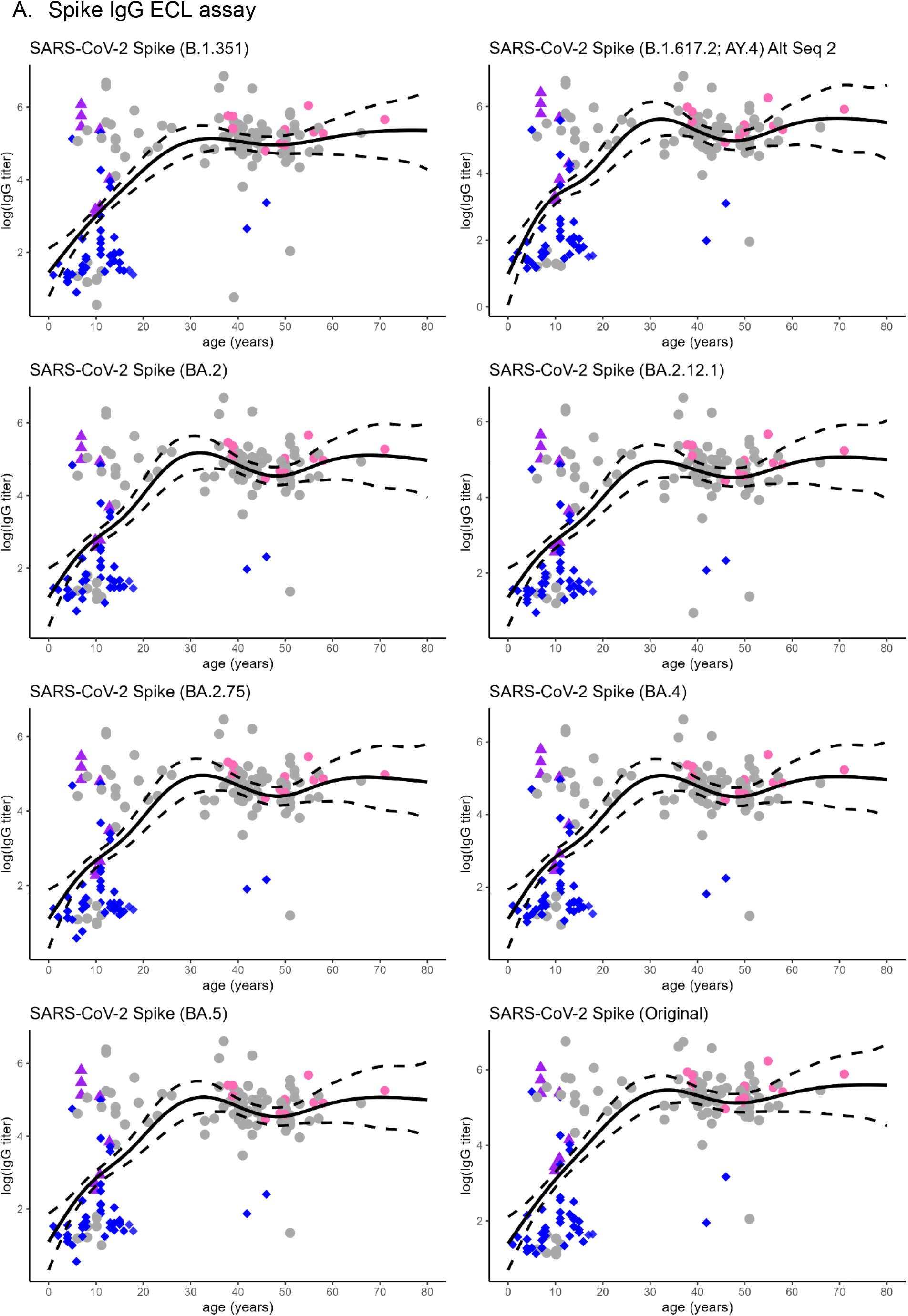
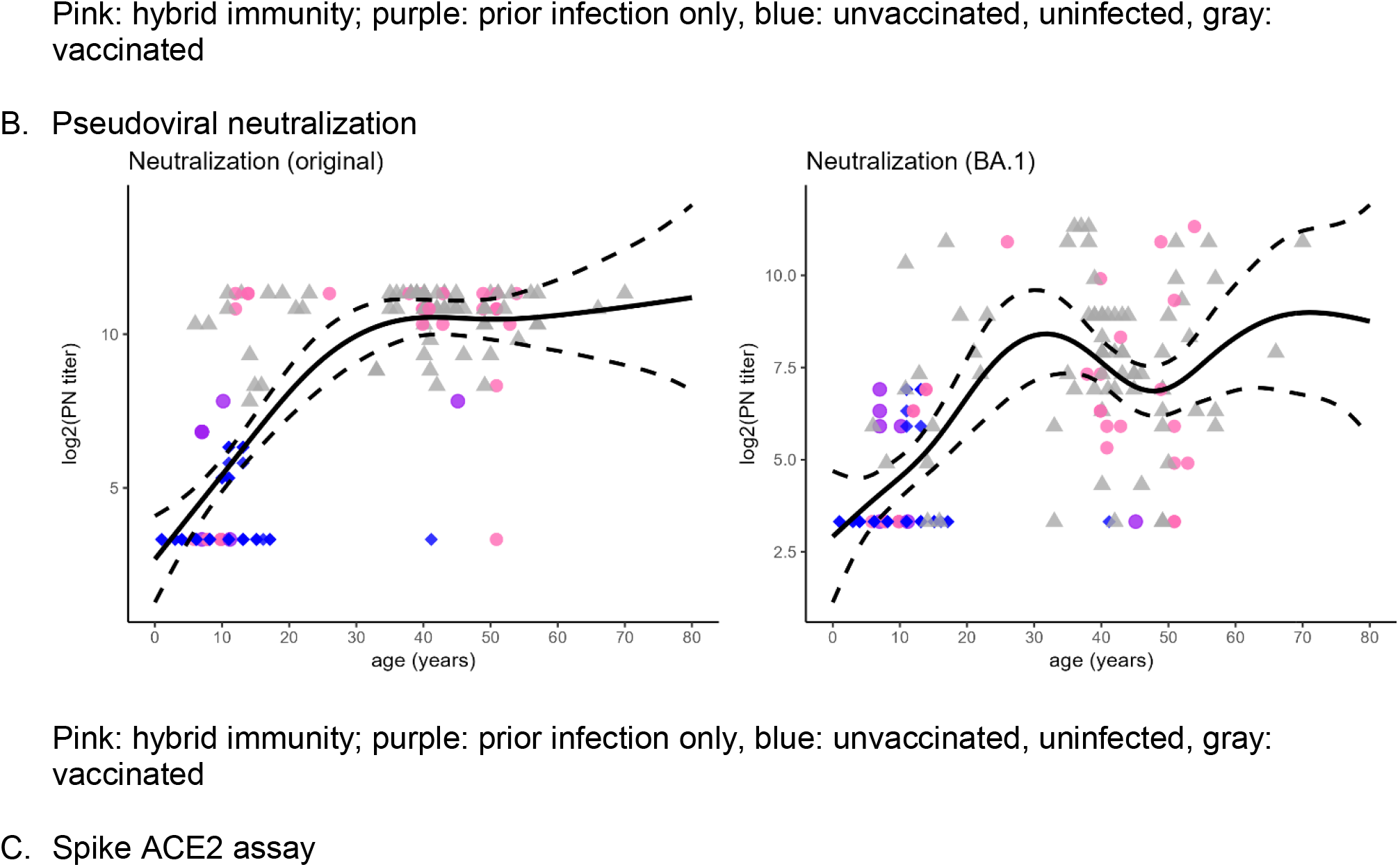
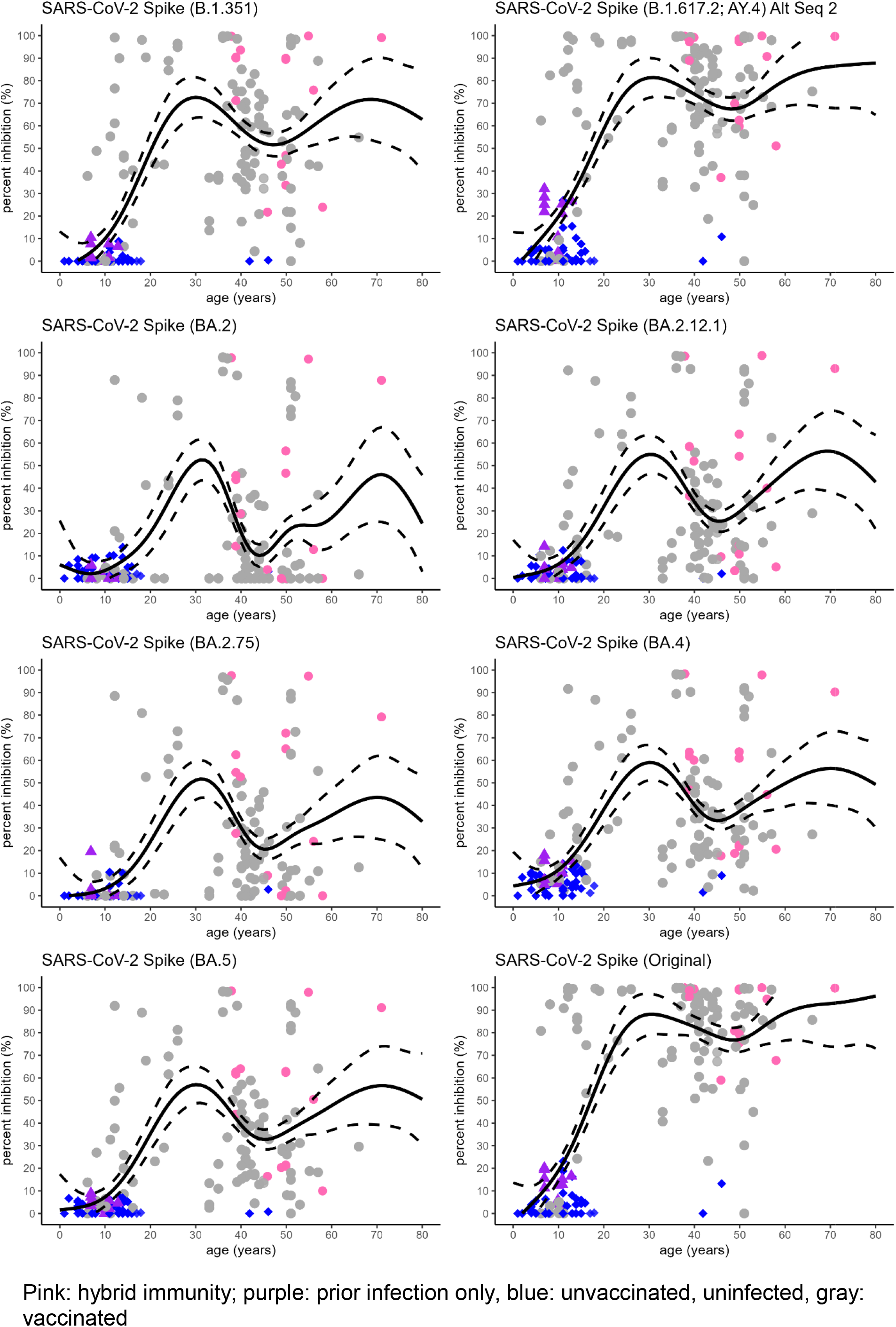
Relationship between age and antibody levels by antigen.

